# Factors Associated with Mental Health Outcomes in Oman during COVID19: Frontline vs Non-frontline Healthcare Workers

**DOI:** 10.1101/2020.06.23.20138032

**Authors:** Muna Alshekaili, Walid Hassan, Nazik Al-Said, Fatima alsulimani, Satish Kumar, Adhra Al-Mawali, Moon Fai Chan, Sangeetha Mahadevan, Samir Al-Adawi

## Abstract

**OBJECTIVE:** This study aims to assess and compare demographic and psychological factors and sleep status of frontline HCWs in relation to non-frontline HCWs

*DESIGN, SETTINGS, AND PARTICIPANTS:* This cross-sectional study was conducted using an online survey from the 8^th^ to the 17^th^ of April 2020 across varied health care settings in Oman accruing 1139 HCWS.

*MAIN OUTCOMES AND MEASURES:* Mental health status was assessed using *Depression, Anxiety, and Stress Scales* (DASS-21), and insomnia was evaluated by the *Insomnia Severity Index* (ISI). Samples were categorized into the frontline and non-frontline groups. Chi-square, odds ratio, and independent t-tests were used to compare groups by demographic and mental health outcomes.

**Results:** This study included 1139 HCWs working in Oman. There was a total of 368 (32.3%), 388 (34.1%), 271 (23.8%), and 211 (18.5%) respondents reported to have depression, anxiety, stress, and insomnia, respectively while working during the pandemic period. HCWs in the frontline group were 1.4 times more likely to have anxiety (OR=1.401, p=0.007) and stress (OR=1.404, p=0.015) as compared to those working in the non-frontline group. On indices of sleep-wake cycles, HCWs in the frontline group were 1.37 times more likely to report insomnia (OR=1.377, p=0.037) when compared to those working in the non-frontline group. No significant differences in depression status between workers in the frontline and non-frontline groups were found (p=0.181).

**CONCLUSIONS AND RELEVANCE:** To our knowledge, this is the first study to explore the differential impacts of the COVID-19 pandemic on different grades of HCWs. This study suggests that frontline HCWs are disproportionally affected compared to non-frontline HCWs. The problem with managing sleep-wake cycles and anxiety symptoms were highly endorsed among frontline HCWs. As psychosocial interventions are likely to be constrained owing to the pandemic, mental health care must first be directed to frontline HCWs.

**Article Summary**

**Methods**

- The study accrued 1139 participants of which 574 were working as frontline HCWs (565 non-frontline workers) serving patients with COVID-19 in different categories of healthcare settings in Oman.
- The following tools used were used alongside the collection of demographic information: The depression, Anxiety and Stress Scale (DASS-21) and Insomnia Severity Index.
- Strengths: This nationally representative study is the first of its kind to investigate the differences in magnitude and the covariates of stress and distress between frontline and non-frontline healthcare workers in Oman.
- Limitations: The use of an online survey and the use of symptom checklists (DASS, ISI) which are typically no match for the ‘gold-standard’ interviews.
- It is also not clear whether the observed mental health outcomes constitute adjustment disorders/ acute stress reaction or present a chronic-type and thus irreversible psychological distress.

## INTRODUCTION

COVID-19, a new strain among the class of corona-virus, has been reported to have first manifested in humans in December 2019, subsequently triggering a global pandemic [1]. Among the countries affected, specifically in the Arabian Gulf, is Oman. On February 24^th^, 2020, Oman reported its first two cases testing positive for COVID-19. The initial report implicated the spread of COVID-19 in Oman via citizens who had travelled abroad [2]. More recently, the Ministry of Health has reported an increasing number of people being diagnosed with COVID-19 with a few deaths and multiple recoveries [3]. With the ever-growing number of confirmed and suspected cases, the workload of healthcare workers (HCWs) has been overwhelming. The long and irregular hours of such continuous and heavy volumes of work have the potential to trigger stress and distress.

Empirical evidence suggests that stress associated with a period of tribulation tends to weaken the immune system, further increasing the risk of diseases [4]. Given this fact, in addition to having a high risk of contracting COVID-19, partly attributed to suboptimal protection [5,6], HCWs are prone to poor mental health outcomes [7,8]. Therefore, early detection among HCWs has the potential to ‘pre-empt’ the development of intransigent, and an advanced pathology of mental health outcomes, thereby helping to reduce the less desirable trend of having compromised HCWs during a pandemic.

The prevalence of stress and distress during times of great tribulation and seismic political, social and economic situations have been extensively investigated [9]. Studies have shown a significant peak of poor coping, maladjustment and the development of emotional disorders in the wake of such unpredictable times [10]. With the current global pandemic of COVID-19, Holmes et al. [11] have emphasized the importance of giving priority to all three tiers of social, psychological, and biological health. As stress and distress have commonly been reported among healthcare workers, often outshining the rate observed in the general population [12,13,14], the question remains whether there are differences in magnitude and the covariates of stress and distress among those working on the frontlines and those who are not. This hypothesis has received scant attention.

While impressionistic reports on the psychosocial issues among healthcare workers have emerged in Oman [8], there is the dearth of studies that address these issues among a nationally-representative sample of healthcare workers. This study from Oman aims to fill this gap in the existing literature. Thus, this study assessed and compared the demographic and psychological factors and sleep status of frontline HCWs vs non-frontline HCWs. Understanding demographic factors that have the potential to tamper with relevant preventative measures and knowing if their magnitude is higher among frontline HCWs will help inform the urgent mechanisms that are needed to preserve the wellbeing and resilience of such subtypes of HCWs [11].

## Methods

### SETTING AND PARTICIPANTS

This cross-sectional study was conducted from 8th April to 17th April 2020 across the varied health care settings in the country. Oman has a universal free healthcare system and is divided into primary, secondary, tertiary, and polyclinics [15]. According to the Ministry of Health of Oman, the first point of contact with healthcare is the primary healthcare setting. If the service seeker should require secondary or tertiary care, then they are referred or transported to the relevant catchment areas with secondary or tertiary care services.

With persisting social distancing, the study proforma was disseminated using emails of representative HCWs working in different regions of the country [16]. After initial contact, potential respondents were also asked to disseminate the information to their colleagues to increase the response rate.

Oman has eleven administrative regions known as governorates or *muhafazah* [16]. Concerted efforts were made to accrue participants from all such regions in the country. One relevant clinical department was randomly sampled from each chosen healthcare setting, and all HCWs in this department were asked to participate in this study. The required sample size corresponding to an acceptable margin of error for proportion (0.1) was calculated. The proportion of HCWs with psychological comorbidity was estimated at 35%, based on an earlier SARS and COVID-19 outbreak report [7, 17]. To allow for analysis of the relevant subgroups, the investigators of this study increased the sample size by 50 percent intending to reach at least 1070 participants. The study proforma was available in both Arabic and English and could be accessed via an online platform (google document) and any information about this study was in the form itself. All respondents provided informed consent. At the end of the study survey, 1160 healthcare workers returned a fully completed study proforma

### Outcomes and Covariates

#### The depression, Anxiety and Stress Scale (DASS-21)

DASS-21 is a self-report screening checklist designed to measure the negative feelings that are broadly categorized as depressive symptoms, anxiety, and stress [18]. Both the English and non-English (including Arabic) versions of DASS-21 have been found to have adequate internal consistency (Cronbach’s alpha scores of > 0.7) [18, 19]. DASS-21 has also been used in Oman and reported to have adequate Cronbach’s α for the three subscales. The present study used the following cut-offs: Depression >= 10; Anxiety >= 8; Stress >=16 [19].

#### Insomnia Severity Index

Insomnia Severity Index (ISI) is a 7-item self-report questionnaire tapping into the severity of insomnia [20]. Both English and non-English including Arabic versions of the ISI have been found to have adequate internal reliability (Cronbach’s alpha scores of > 0.7 [21]. A 5-point Likert scale was used to rate each item (e.g., 0 = no problem; 4 = very severe problem), yielding a total score ranging from 0 to 28. A previous study suggested that a cut-off score of 14 was deemed adequate for detecting clinical insomnia with a sensitivity of 82.4% and specificity of 82.1% [22].

#### Demographic factors

The study proforma included socio-demographic data (nationality, gender, age, marital status), type of medical setting (primary, secondary, tertiary care, or polyclinic) and whether they were directly engaged in clinical activities such as diagnosing, treating, or providing nursing care to patients with elevated temperatures or patients with confirmed Covid-19 infection. Those who responded as diagnosing, treating, or providing nursing care were identified as ‘frontline HCWs’. Those participants who had no contact with the units assigned to handle services for COVID-19 patients were defined to constitute second-line workers or ‘non-frontline HCWs’.

Participants’ job type (physician, nurse, and allied healthcare professional) was also sought. Allied healthcare professions included pharmacists and other medical staffing including laboratory technicians. Finally, the participants were also asked whether they had previously sought consultation for psychiatric disorders (‘yes’ / ‘no’).

#### Ethical Issues

This study adhered to the American Association for Public Opinion Research reporting guidelines [23]. Ethical approval was obtained before the commencement of the study from the local IRB, Directorate General of Planning and Studies, Centre of Studies and Research, Ministry of Health (MOH/ DGPS/CSR/20/2311). Written consent was sought from participants and they were told specifically that their involvement could be terminated if they wish so without undue consequences. The survey was anonymous, and confidentiality of information was assured.

#### Statistical Analysis

Data analysis was performed using SPSS statistical software version 23.0 (IBM Corp). Descriptive statistics were used to explore the profile of the samples in terms of their demographic and psychological outcomes. Samples were categorized into the frontline and non-frontline groups. Chi-square, odds ratio, and independent t-tests were used to compare groups by demographic and psychical factors. All significant tests were set at 5% alpha level.

## Results

This cross-sectional study was conducted using an online survey from the 8^th^ to the 17^th^ of April in 2020 across different health care services in Oman. In total, we received 1167 questionnaires of which 28 were determined to be incomplete on examination. Thus, we only included 1139 records for further analysis.

### Demographic and psychological outcomes of the study samples

In Table 1, among the 1139 HCWs, 228 (20.0%) are males, and 911 (80.0%) are females. Their average age was 36.3 ± 6.5 (Mean ± SD) and range from 21 to 65 years. The majority are Omani (n=981, 86.1%) and are married (n=987, 86.9%). A total of 574 (50.4%) were directly involved in diagnosing, treating, and taking care of confirmed or suspected cases of COVID19 (frontline group). There were 390 (34.2%), 164 (14.4%), 478 (42.0%), and 106 (9.3%) were working in primary, secondary health care, tertiary health care as well as polyclinics respectively. Among those HCWs, 384 (33.7%), 449 (39.5%), and 305 (26.8) were physicians, nurses, and allied health profession, respectively. Concerning psychological outcomes, 368 (32.3%), 388 (34.1%), 271 (23.8%), and 211 (18.5%) respondents reported symptoms of depression, anxiety, stress, and insomnia, respectively while working during the pandemic period.

**Table 1.**
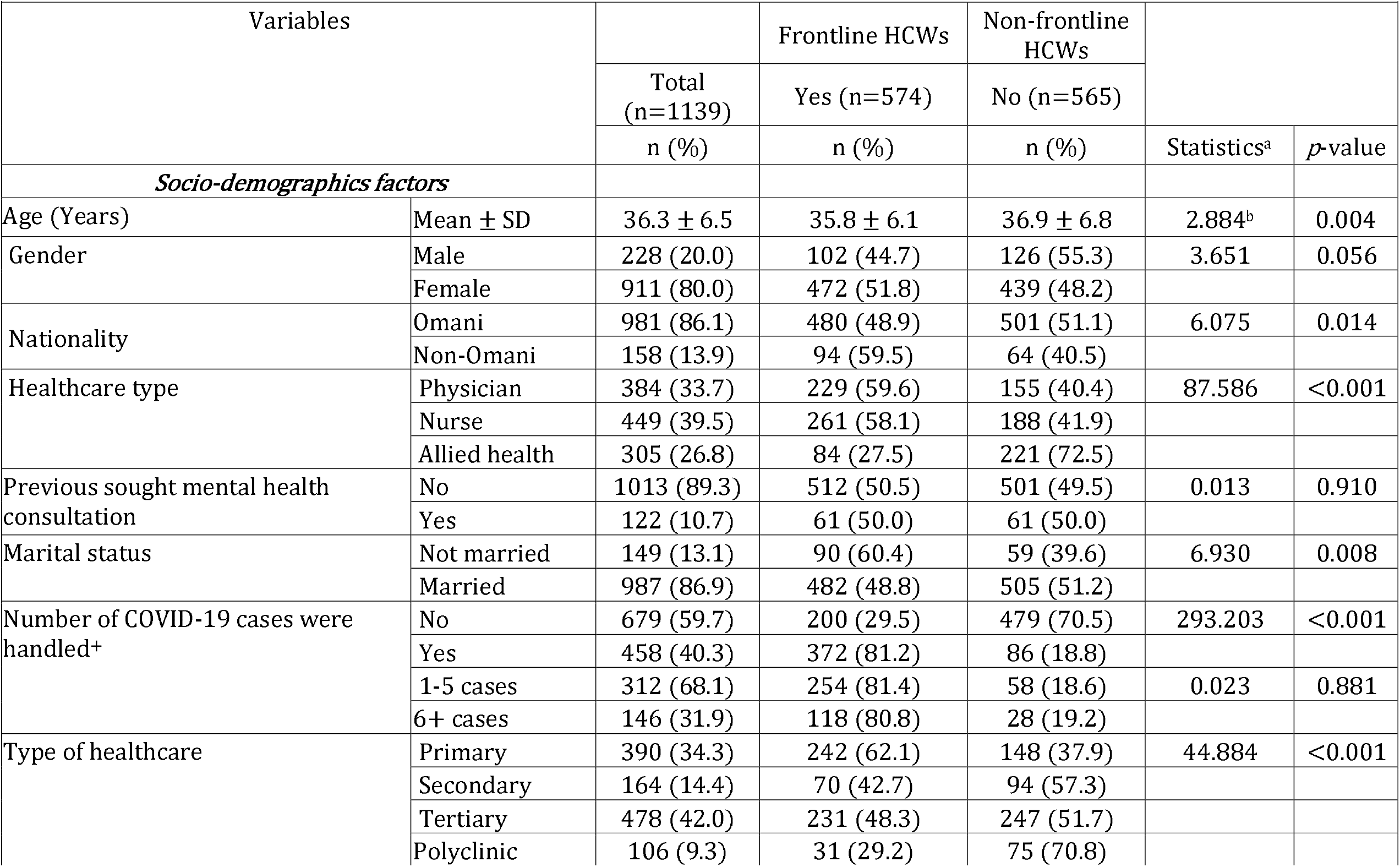
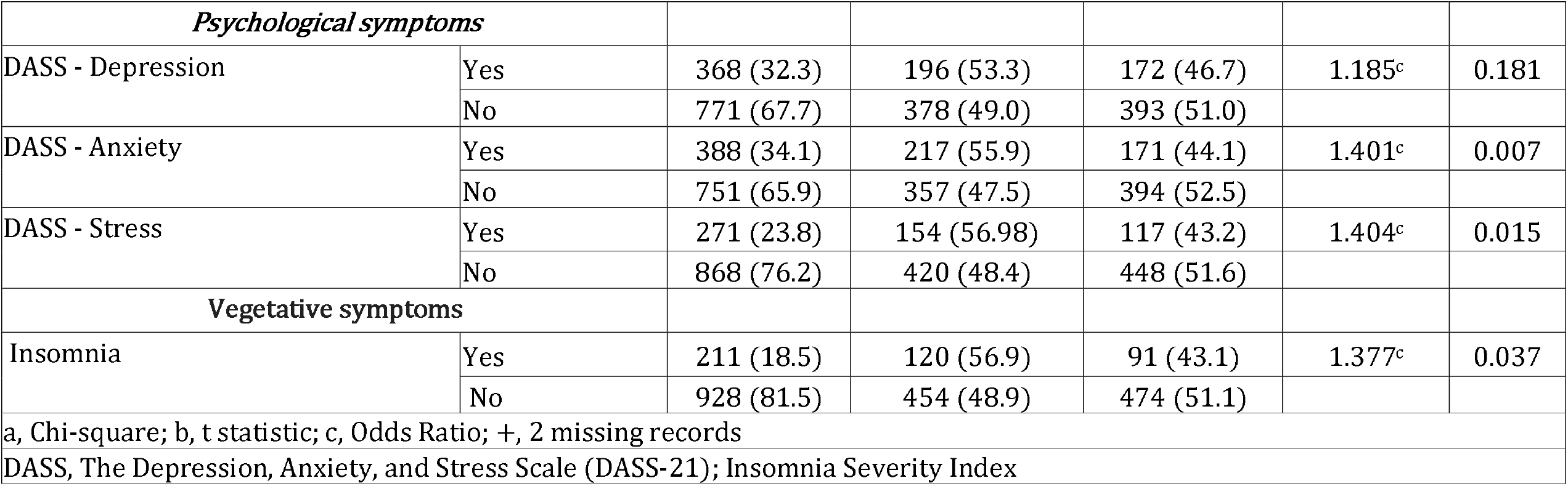
Comparison of the frontline with non-frontline staff in association of demographic and psychological factors during the impacts of COVID-19 in Oman.

### Comparison of frontline and non-frontline staff on demographic and psychological outcomes

In Table 1, significant differences were found between the two presently defined cohorts of HCWs - frontline and non-frontline groups. The frontline group comprised of members younger in age (36.3 ± 6.5, p=0.004) with more of them being non-Omani (n=94, 59.5%, p=0.014), physicians and nurses (n=490, 58.8%, p<.001), not married (n=90, 60.4%, p=0.008), handled COVID-19 cases (n=372, 81.2%, p<.001), and working in primary health care setting (n=242, 62.1%, p<.001) as compared to the non-frontline group. With regard to psychological outcomes, members of the frontline group were 1.4 times more likely to have anxiety (OR=1.401, p=0.007) and stress (OR=1.404, p=0.015) as compared to the non-frontline group. In considering insomnia, the frontline group was 1.37 times more likely to experience the same (OR=1.377, p=0.037) as compared to the non-frontline group. No significant differences in depression status were found between the two groups (p=0.181).

## Discussion

Various mechanisms to come to grip with the COVID-19 pandemic, including travel restrictions, quarantines, and curfews which, in turn, have severely disrupted the social and economic activities of the society, nation, or for that matter the world [24], have been proposed. While the impact of socio-economic activities due to COVID-19 has been widely acknowledged in the countries of the Arabian Gulf [25], what has been overlooked is the fact that healthcare workers (HCWs) are in the frontline in the COVID-19 pandemic which, in turn, would suggest the importance of examining their resilience in the light of those challenges.

Being a “once-in-a-century pandemic” [26], some of the misgivings affecting HCWs include the fear of contracting a lethal virus and spreading it to the rest of their social network, lack of evidence-based prevention and intervention, lack of essential protective gear and the fact that the pandemic requires protracted shifts with a high volume of patients with different degrees of pathology and severity [27,28]. This would imply that HCWs are now working in a uniquely hazardous situation and are thus vulnerable to stress and distress. In addition to operational stresses, some preliminary studies have suggested that psychosocial dysfunctions are rife among HCWs [28, 29]. A recent systematic literature review and meta-analysis covering the literature of the pre-pandemic COVID-19 period suggest that 7.0% to 75.2% of HCWs are burned out [30]. This huge discrepancy in the prevalence of burnout hinges on country-specific factors, applied instruments and cut-off-criteria for burnout symptomatology [31]. The prevalence of burnout among HCWs appears to outstrip the general population [32]. Similarly, in addition to burnout, the prevalence of depressive symptoms, anxiety, and stress among HCWs are also higher than the general population [12,13,14]. However, since the higher level of stress and distress among HCWs as compared to the general population has been a trend existing even before the pandemic, it not clear whether the emerging high level of mental health outcomes owes its onset to COVID-19. One approach to disentangling this issue is to compare mental health outcomes between frontline HCWs vs non-frontline HCWs. This study had therefore embarked on the assessment and comparison of demographic and psychological factors and sleep status of frontline versus non-frontline HCWs.

The present study accrued 1139 HCWs from different parts of the country. As the HCWs in Oman are predominantly female [33], this study is in line with the observed ‘effeminization’ of healthcare as 80.0% of the present participants were female. Approximately 50% fulfil the present definition of ‘frontline HCWs’ who, in their clinical practices, diagnose, treat, and take care of confirmed or suspected cases of COVID-19 in their respective clinics across the country. The cohort consisted of physicians, nurses, and allied health professionals.

To tap into the levels of depression, anxiety, and stress, the *Depression, Anxiety, and Stress Scale* (DASS-21) was used. Of the present cohort, comprised of both frontline and non-frontline HCWs, 32.3% endorsed case-ness for depressive symptoms, 34.1% for anxiety, and 23.8% for stress. In Singapore among HCWs using DASS-21, Tan et al. [34] have reported 8.9% case-ness for depression, 14.5% for anxiety, and 6.6% for stress. Using different screening tools, Lai et al. [7] have reported 50.4%, 44.6%, and 71.5% symptoms of depression, anxiety, and distress respectively among HCWs in Hubei province in China. Lai’s study indicated that 34.0% of their sample had an elevated score of insomnia which appears to be lower compared to the prevalence of 38·9% among HCWs investigated as part of the studies included in their systematic review and meta-analysis [29]. Putting these studies together and within the background of the general population, other than the lower rate of depression in Singapore, the magnitude of mental health outcomes appears to be higher among HCWs when compared to the general population. In the general population, the prevalence of depression, anxiety, and insomnia have been estimated to be 11.1% [12], 5.3%, 7.3% [13], and 10% - 30% [14] respectively. Low mental health outcomes among HCWs in Oman and Singapore could be attributed to the preparedness phase the country underwent as the first cases were registered much later than when the World Health organization declared COVID-19 a global pandemic [1]. While studies on the status of mental health outcomes and sleep status have been forthcoming from different parts of the world, many of them are single-center [8] and regional studies [7] with some of the catchment areas not being defined [34]. A study with a nationally representative sample of HWCs taking into account both the frontline and non-frontline are therefore warranted.

The second aim of the present study was to compare demographic and psychological outcomes among frontline and non-frontline HCWs. The present data suggest that frontline HCWs are likely to be younger, single, physicians or nurses working in primary healthcare and are required to handle COVID-19 cases. The majority of frontline HCWs were non-Omani, a trend that is worth contemplating. Despite the effort to ‘Omanize’ the healthcare infrastructure, foreign nationals still form the bulk of HCWs in Oman [35]. The COVID-19 pandemic has resulted in travel restrictions and an expected economic recession, resultant job security, and being cut off from their country of origin for the migrant population [36]. It remains to be seen whether these factors have rendered non-Omani HCWS to be more vulnerable to the presently observed mental health outcomes.

In psychological outcomes, compared to non-frontline HCWs, frontline HCWs were more likely to endorse anxiety symptoms and stress. A similar trend was observed with insomnia. Interestingly, the depressive symptoms did not emerge as being significant in the equation employed to differentiate between frontline vs non-frontline HCWs. Oman is characterized by a collectivistic society that is in direct contrast to western individualistic societies [37]. In such a society, anxiety symptoms (‘I experienced trembling in the hands’) and stress (‘I felt that I was using a lot of nervous energy’) tend to be perceived to be a veneer of physical symptoms and are therefore likely to be endorsed. In contrast, depressive symptoms (‘I felt down-hearted and blue’) are thought to be more of a weakness of character than a manifestation of ‘disease’. As psychological outcomes are increasingly recognized to emerge as a consequence of COVID-19 [11], more studies are needed to decipher the culturally-specific idioms of distress intimately tied to mental health outcomes during the pandemic.

### Limitations

Most psychosocial studies of this nature tend to have many limitations owing to the amorphous variables under scrutiny. Firstly, conducting a national wide survey requires proper logistics which was not feasible during the lockdown. An online survey is known to marred by the fact that it tends to accrue a selective population who are technologically savvy and more familiar with the evolving ‘internet culture’ [38]. Notwithstanding such a view, this study appears to have reached its targeted population based on the estimated sample size. Oman has established that > 71% of the total population (4.6 million) has access to internet services [38]. Secondly, DASS-21 and ISI are no match for the ‘gold-standard’ interviews such as those that follow the *Diagnostic and Statistical Manual of Mental Disorders* and World Health Organization *Composite International Diagnostic Interview* (CIDI). However, quick symptom checklists such as DAS-21 and ISI are the only viable tools to conduct such a study given the current circumstances [7]. Lastly, time factors are also considered important for quantifying the presence of psychological disorders. Within this view, it not clear whether the observed mental health outcomes constitute adjustment disorders/ acute stress reaction or present a chronic-type and thus irreversible psychological distress. Follow-up studies in this regard are therefore warranted.

## Conclusion

COVID-19, a new strain among the class of Coronavirus, has recently gripped all corners of the world triggering a global public health emergency. Within the background of high rates of poor coping among HCWs even before the pandemic, studies are needed to explore how frontline HCWs fare compared to non-frontline HCWs in this regard. This study highlighted and appeared to be congruent with other studies in suggesting that the COVID-19 outbreak has triggered a higher rate of depressive symptoms, anxiety, and insomnia among HCWs. In comparing frontline and non-frontline HCWs, the present data suggested that frontline HCWs were likely to be younger non-Omani physicians or nurses who were single, and working in primary healthcare. It is therefore paramount to offer timely psychological intervention for the HCWs to promote coping and resilience among these vulnerable HCWs.

## Data Availability

All data generated and analysed during this study are included as part of this article.

## ARTICLE INFORMATION

### Corresponding Authors

Dr. Samir Al-Adawi, Department of Behavioral Medicine, College of Medicine & Health Sciences, Sultan Qaboos University, P.O. Box 35, Al Khoudh 123, Muscat, Oman

Al Masarra Hospital, Ministry of Health, Wilayat Al Amerat, Muscat (**Alshekaili M**., **Hassan W**., **Al-Said N**., **alsulimani F**.); Centre of Studies & Research, Directorate General Planning and studies, Ministry of Health, Oman (**Kumar S**., **Al-Mawali A**.); Department of Family Medicine & Public Health, Sultan Qaboos University, Muscat, Oman(**Chan, Moon Fai**); Department of Behavioral Medicine, Sultan Qaboos University Hospital, Muscat, Oman (Mahadevan **S**., **Al-Adawi S**.)

## Author Contributions

Drs Muna Alshekaili, Walid Hassan, Nazik Al-Said, Fatima alsulimani and Satish Kumar had full access to all of the data in the study and take responsibility for the integrity of the data and the accuracy of the data analysis.

### Concept and design

Muna Alshekaili, Samir Al-Adawi.

### Acquisition, analysis, or interpretation of data

Muna Alshekaili, Walid Hassan, Satish Kumar, Adhra Al-Mawali,, Moon Fai Chan

### Drafting of the manuscript

Muna Alshekaili, Walid Hassan, Samir Al-Adawi.

### Critical revision of the manuscript for important intellectual content

Samir Al-Adawi, Sangeetha Mahadevan, Moon Fai Chan

### Statistical analysis

Walid Hassan, Moon Fai Chan

### Administrative, technical, or material support

Satish Kumar, Adhra Al-Mawali,

### Supervision

Muna Alshekaili, Walid Hassan.

## Conflict of Interest Disclosures

None reported.

### Funding/Support

This research received no specific grant from any funding agency in the public, commercial or not-for-profit sectors.

### Additional Contributions

We thank all the participants who contributed to our work.

### Data Statement

All data generated and analysed during this study are included as part of this article.

## Notes

### Competing Interest Statement

The authors have declared no competing interest.

### Author Declarations

Ethical approval was obtained before the commencement of the study from the local IRB, Directorate General of Planning and Studies, Centre of Studies and Research, Ministry of Health (MOH/ DGPS/CSR/20/2311)

